# The agrarian imperative as the foundation for how farmers and ranchers diagnosed with alpha-gal syndrome find meaning in their experience: A mixed methods approach

**DOI:** 10.64898/2026.10.29.26366631

**Authors:** Alexandrea M. Welch, Cheryl L. Beseler, Shaun T. Cross

## Abstract

The agrarian imperative is the drive of agricultural workers to use their land to provide for their families and communities, even when the environment is hazardous and success is often out of their control. Alpha-gal syndrome (AGS), an emerging tickborne allergy to mammalian derived products, is challenging the agrarian imperative by impacting the worker’s ability to maintain their operation. This study aimed to characterize farmers and ranchers’ experiences after receiving an AGS diagnosis related to their land and their work. Using the framework of the agrarian imperative theory and a mixed methods approach, we aimed to determine the impact on daily work tasks, the wellbeing burden of the diagnosis, the financial challenges of the operation and daily life, and dietary changes. Survey data from 201 farmers and ranchers diagnosed with AGS were analyzed alongside qualitative data from interviews with 14 participants. The respondents felt as though their wellbeing was affected, and felt distressed, depressed, and anxious. Anxiety was the most frequently reported mental health impact of AGS due to forced dietary changes, but distress was more highly associated with work factors. Nearly half of the sample were experiencing distress due to having to modify their work tasks and worrying about the productivity of the agricultural operation. We found agricultural operators with animals are more likely to experience greater work challenges. Maintaining a productive farm was the most important objective and reflects the agrarian imperative, but additional costs associated with AGS complicates their work and life.

## Introduction

Farmers and ranchers experience significant occupational stress, exacerbated by the unique demands of agricultural work [1–3]. In many agricultural communities a high percentage of physicians’ visits are due to stress-related illnesses, and farming is one of the most stressful occupations [4]. Key occupational risk factors contributing to stress among farmers and ranchers include input costs, commodity prices, weather events, pesticide exposure, injury risks, isolation, role conflict, and time pressure [1–3]. Larger farms with greater than $100,000 in annual farm income and farmers younger than 40 are more vulnerable to financial stressors due to an increase in debt and limited fiscal capacity to meet their obligations [5, 6]. Unlike other professions, farmers and ranchers who raise animals are often unable to take time off if an injury or illness occurs; vacations are rare [7].

The foundation of the agrarian imperative is the motivation of agricultural workers to use their land sustainably to provide for their families and communities for present and future generations. The agrarian imperative is the rationale behind agricultural workers’ drive to persevere when their work environment is hazardous and success is often out of their control [8]. Despite the aforementioned challenges, farmers and ranchers often continue to work due to their commitment to their operations, families, and communities leading to the idea that the agrarian imperative is an innate trait in generational farm families. Agricultural operators have a deep connection to the land that they cultivate and they find fulfillment in their farming lifestyle. They thrive living close to animals and nature and experience self-determination that is tightly linked to their wellbeing [9].

However, an emerging tickborne disease is challenging the agrarian imperative. Alpha-gal syndrome (AGS) is an immunoglobulin E (IgE)-mediated allergy to mammalian derived products [10]. AGS can trigger allergic reactions including urticaria, angioedema, anaphylaxis, and/or gastrointestinal symptoms including diarrhea, abdominal pain, nausea, and vomiting [11], and reactions are typically delayed 2-6 hours after consuming mammalian byproducts [12]. Studies have shown individuals living and working in rural environments, such as farmers and ranchers, are at a greater risk of tick exposure, including lone star ticks which are associated with AGS [13]. Furthermore, because AGS causes a reaction to mammalian byproducts, this poses a threat to farmers and ranchers who work with mammals and their products daily and may react to unique sources of alpha-gal such as amniotic fluids [14]. There is a critical need to understand the work impacts and associated mental health effects of an AGS diagnosis on farmers and ranchers who are in tick habitats, consume mammalian products, and work with livestock daily.

In a previous study of 201 individuals diagnosed with alpha-gal syndrome, we identified four distinct symptom clusters [15]. The most populated class was the gastrointestinal group which was 31.8% of the sample. The smallest class were those who were labeled the most severe group because they reported frequently having a range of symptoms and were 16.4% of the sample. The groups with mild symptoms (infrequent but a broad range of symptom types) or allergy-type symptoms were each 25.9% of the sample. This wide-ranging symptomatology can make diagnosis a challenge [16].

This study aimed to characterize farmers and ranchers’ experiences after receiving an AGS diagnosis related to their land and their work. Using the framework of the agrarian imperative theory and a mixed methods approach, we aimed to determine the impact on daily work tasks, the wellbeing burden of the diagnosis, the financial challenges of the operation and daily life, and dietary changes. In agrarian populations, mental health, work, and diet is interconnected to the land and the generational commitment to the farm or ranch they raise plants and animals on.

## Methods

### Participants and recruitment

The original quantitative survey was administered through a Facebook AGS group, though subsequent sharing by others and organizations occurred from this initial posting. The qualitative sample was a convenience sample recruited from within the quantitative survey conducted to obtain information from those who had been diagnosed with AGS. Survey participants were asked if they would be willing to be contacted for personal interviews. Those who acquiesced to being contacted again were invited to participate in an online video interview. We stratified participants by age and sex, and individuals were randomly invited with equal representation across each stratum. Contacting of 57 potential participants resulted in 14 completed interviews. Participants were provided a $50 gift card to compensate them for their time. Interviews were 45 to 60 minutes in length and conducted online using Zoom. This work was approved by the University of Nebraska Medical Center Institutional Review Board (IRB) 0562-25-EX.

### Measures: quantitative data

The primary goal of this study was to understand the impact of an AGS diagnosis on work practices and behavioral wellbeing. Agricultural operators were asked whether their diagnoses required them to modify their agricultural tasks, whether certain tasks caused higher risk for experiencing symptoms than others, and whether these challenges caused them to worry about their work productivity. We also asked whether symptoms affected wellbeing, or caused distress, depression, anxiety, or feelings of hopelessness. We specifically asked whether making food choices caused anxiety, whether respondents were confident in their ability to manage their condition, and whether they had a support system. We also assessed whether previously published AGS symptom classes were associated with work variables and measures of wellbeing [15].

### Measures: qualitative data

As the quantitative survey data alone could not provide full insights and experiences of those affected, we performed a qualitative study comprised of 14 one-on-one interviews. We broadly asked about a range of topics using the quantitative questions as a guide. We asked about a family history of allergies or autoimmune disorders, history of exposure to tick bites, modifications to work tasks and exposures, economic impacts due to both farm income fluctuations and alterations in diet and products used, impacts on social and family relationships, and mental health reactions to the diagnosis. The questions remained in a generalized format to allow respondents to disclose what they felt was relevant to them.

### Analysis: quantitative

Descriptive statistics were used to summarize the binary (no=0, yes=1) responses to the quantitative work-related factors and mental wellbeing measures. We used the previously identified groups from a latent class analysis [15] and tested with a chi-square statistics whether symptom groups were associated with the mental health impacts assuming that greater severity of symptoms would increase impacts. We used chi-square tests to assess whether the work-related variables (modifying work, risk of farm work, having help with farm work, and productivity worries) were related to the mental health variables (wellbeing, distress, depression, anxiety, and hopelessness). We created four categories from the variables for having had to modify work tasks and expressing worries about farm productivity: (1) no modification nor worries; (2) modifications but no worries; (3) no modifications and worries; and (4) both modifications and worries. The newly coded four category explanatory variable was used as three indicator variables in logistic regression models with the outcomes being separate models for the binary variables feeling distress, feeling depressed, feeling anxious, or feeling hopeless. The reference group was comprised of those with no work modifications made and no productivity worries. All analyses were conducted using SAS version 9.4 [17]. Results were considered statistically significant at p<0.05. Graphs were created in the R package ggplot2 [18].

### Analysis: qualitative

The purpose of this phenomenological study is to describe the mental health impacts of an AGS diagnosis in farmers and ranchers. The analysis is based on the agrarian imperative theory [8], a hypothesized psychological construct. With a theory-based analysis, the qualitative analysis used a deductive, directed content analysis followed by taking a phenomenological perspective by summarizing the commonly expressed meaning of respondents’ experiences. We aimed to capture the internal experiences that respondents reported after their diagnosis and how these experiences are based in their connection to their land and their work.

We used a predefined coding scheme and did a directed content analysis [19,20]. Although we had the major themes from theory and the quantitative study, we aimed to identify subthemes based on the transcripts. Each transcript was read thoroughly by two coders and was annotated by highlighting references to the phenomenon of AGS. Subthemes were identified in the first five transcripts and further expanded upon when analyzing additional transcripts. No new codes were identified after about ten transcripts and saturation was reached. The two coders (AW and CB) met twice to reach a consensus on the coding. The subthemes were clearly identified and there was no disagreement. The authors then characterized the experiences based on the subthemes in the context of the agrarian imperative. The meaning of the AGS experience was hypothesized to be shared across participants but was expected to differ only by the intensity with which the participant was engaged in agricultural activities and the severity of their AGS symptoms. Many farmers work full-time on their operations and others part-time.

### Analysis: Mixed Methods

An Explanatory Sequential Design [21] was used in this study with the quantitative analysis first followed by interviews to enlighten the quantitative responses. The mixed methods encompassed two major goals. First, identification of the sources of stress and reported mood disorders in the quantitative responses and the primary sources of the perceived stressors. Second, an expanded understanding of how AGS altered farmwork tasks and what modifications were made by participants to complete these tasks. The themes from the qualitative study were matched to the questions in the quantitative survey and assessed for context.

## Results

### Quantitative sample

The AGS cohort was overwhelmingly female (n=149, 74.1%) and the majority operated a ranch (n=127, 63.2%). The majority of respondents endorsed having to modify their work tasks, reported some tasks were riskier than others, worried about productivity of their operations due to their diagnosis, but also reported having someone to help with the farm work. The respondents were also likely to feel their wellbeing was affected, and felt distressed, depressed, and anxious. Less than half reported feeling hopeless and seeking out behavioral healthcare. Anxiety around eating was high. Respondents felt as if they had a good support system and felt confident that they could manage their AGS.

When examining the associations between the work-related variables, we found that modifying work tasks was associated with task-related risks (p<0.0001) and productivity worries (p<0.0001). Modifying work tasks and productivity worries were associated with a number of mental health outcomes (Figure 1). Having to modify work-related tasks was associated with distress (p=0.0001), wellbeing (p=0.007), anxiety (p=0.002), and depression (p=0.02). Productivity worries was associated with distress (p<0.0001) and depression (p=0.03). Managing risky tasks was associated with wellbeing (p<0.0001), anxiety (p=0.0002), and distress (p=0.006) and individuals who endorsed performing risky tasks were more likely to seek behavioral healthcare (p=0.03). There was no statistically significant association between social support and mental health, possibly because 87.6% felt supported (Table 1). Having to modify work tasks and having to do risky tasks were more impactful on mental health than worries about farm productivity. Anxiety was the most frequently reported mental health impact of AGS (85.1%; Table 1), but distress was more highly associated with work factors.

**Figure 1.**
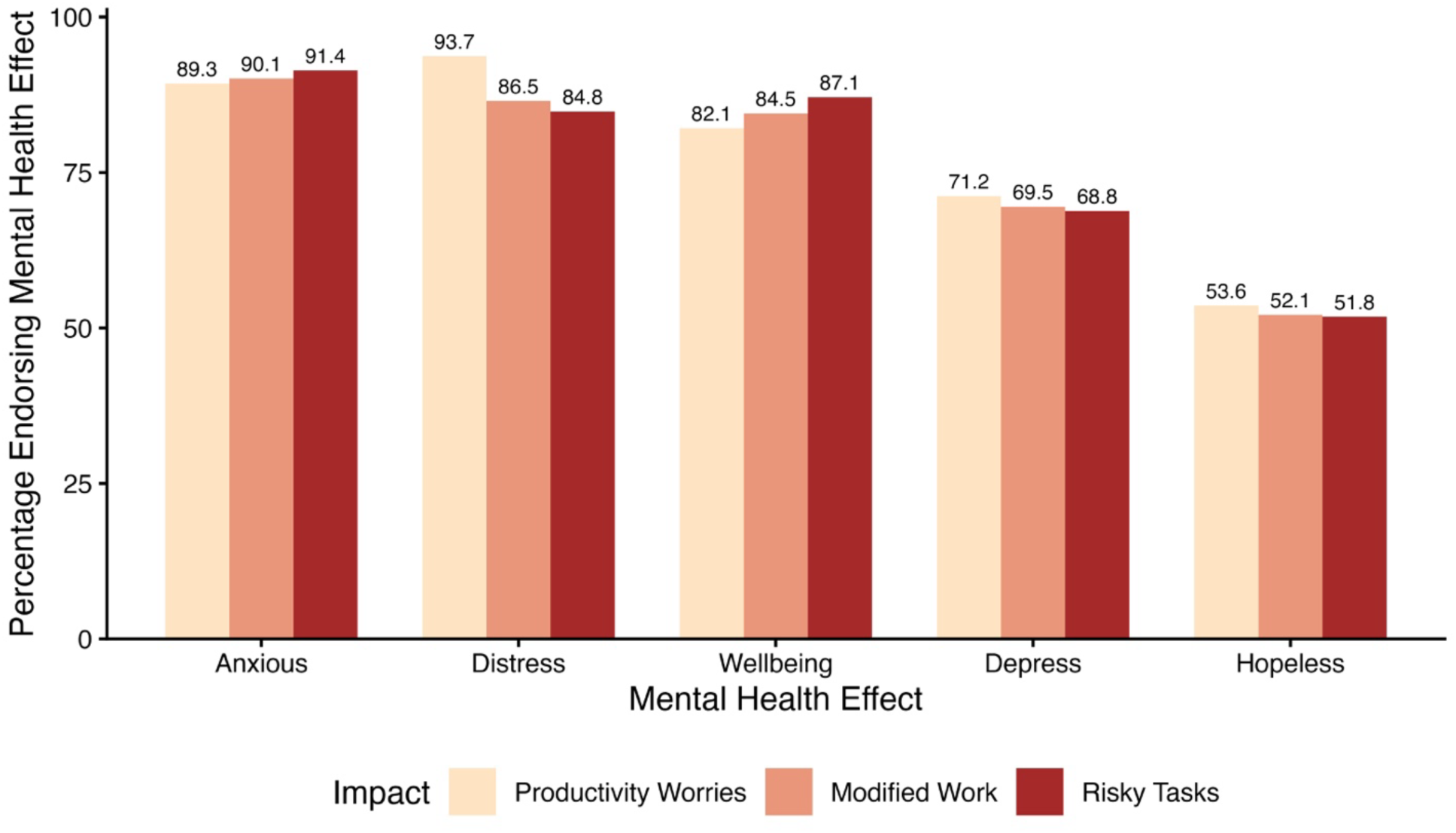
Percentage of AGS respondents who endorsed a mental health impact and being required to modify their work tasks, perform risky tasks, and their worries of operation productivity, 2025.

**Table 1.** Farmwork-related and mental health impacts of 201 respondents diagnosed with AGS, 2025.

| Work and mental health characteristics | n (%) |
| --- | --- |
| Have you had to modify any work-related tasks since the diagnosis? |  |
| Yes | 142 (70.6) |
| No | 59 (29.4) |
| Are there any specific job tasks or exposures that pose a greater risk than others since your alpha-gal diagnosis? |  |
| Yes | 139 (69.2) |
| No | 62 (30.8) |
| Is there anyone who can help you on your farm or ranch? |  |
| Yes | 106 (76.3) |
| No | 33 (23.7) |
| Do you worry about the productivity of your operation because of the alpha-gal diagnosis? |  |
| Yes | 112 (55.7) |
| No | 89 (44.3) |
| Are there any symptoms of alpha-gal that you are experiencing that have affected your well-being? |  |
| Yes | 160 (79.6) |
| No | 41 (20.4) |
| Do you feel like this diagnosis has caused you distress in addition to managing your farm or ranch? |  |
| Yes | 159 (79.5) |
| No | 41 (20.5) |
| Would you say that the diagnosis caused you to feel depressed? |  |
| Yes | 129 (64.5) |
| No | 71 (35.5) |
| Would you say that the diagnosis caused you to feel anxious? |  |
| Yes | 171 (85.1) |
| No | 30 (14.9) |
| Has this diagnosis given you feelings of hopelessness? |  |
| Yes | 96 (47.8) |
| No | 105 (52.2) |
| Have you seen any behavioral health professionals as a result of the diagnosis? |  |
| Yes | 45 (22.4) |
| No | 156 (77.6) |
| Has alpha-gal caused you to feel anxiety in making food choices? |  |
| Yes | 186 (93.0) |
| No | 14 (7.0) |
| Do you feel confident managing this condition? |  |
| Yes | 129 (64.2) |
| No | 72 (35.8) |
| Do you have a good support system? |  |
| Yes | 176 (87.6) |
| No | 25 (12.4) |

Although having a good support system was not associated with modifying work tasks, risky tasks, or productivity worries. Social support was significantly associated with feeling less depressed (p=0.03), feeling less hopeless (p=0.03), and feeling more confident in managing AGS symptoms (p=0.007).

### Differences by sex

In the current study, women reported greater anxiety than men (p=0.001) but there was no differences in reported depression by sex (p=0.52). Men were more likely to report seeing a behavioral health professional than women (p=0.04). Women were more likely to report feeling anxiety about food than men (p=0.0006), possibly because they are usually the family member responsible for meal preparation and might experience symptoms when cooking for the family.

### Work modification and operation productivity

Sex was a confounder only in the anxiety model and was added as a covariate to that model. The type of operation, farm or ranch, was not associated with any other covariates. Respondents who both modified their work tasks and worried about productivity had significantly increased odds of endorsing each of the four mental health impacts compared to those who did not have these combined work factors (Table 2). Nearly half of the sample (n=92, 45.8%) were experiencing distress due to having to modify their work tasks and worrying about the productivity of the agricultural operation. The odds ratio of 18 is twice that of those who did not need to modify their tasks and had no productivity worries. Modifying work tasks alone, without productivity worries, resulted in a significantly elevated odds of distress, depression, and anxiety, but not wellbeing. Worry about the operation’s productivity was significantly associated only with feeling distressed, although the odds ratio was elevated for each of the other mental health impacts.

**Table 2.** The impacts of productivity worries, modifying work tasks and both on mental health in 201 participants diagnosed with AGS, 2025.

| Model | OR (95%CI) |
| --- | --- |
| Wellbeing affected |  |
| Neither modified work tasks nor worried about productivity | Reference |
| Worried about productivity | 0.83 (0.26, 2.59) |
| Modified work tasks | 2.03 (0.75, 5.46) |
| Modified work tasks and worried about productivity | 2.70 (1.10, 6.63) |
| Felt distressed |  |
| Neither modified work tasks nor worried about productivity | Reference |
| Worried about productivity | 9.47 (1.93, 46.5) |
| Modified work tasks | 2.71 (1.12, 6.53) |
| Modified work tasks and worried about productivity | 18.1 (6.03, 54.3) |
| Felt depressed |  |
| Neither modified work tasks nor worried about productivity | Reference |
| Worried about productivity | 3.02 (0.96, 9.50) |
| Modified work tasks | 2.51 (1.06, 5.95) |
| Modified work tasks and worried about productivity | 3.23 (1.48, 7.05) |
| Felt anxious |  |
| Neither modified work tasks nor worried about productivity | Reference |
| Worried about productivity | 2.63 (0.66, 10.6) |
| Modified work tasks | 3.96 (1.25, 12.6) |
| Modified work tasks and worried about productivity | 6.73 (2.28, 19.9) |

### Associations with class membership

Class membership was associated with having to modify work tasks (p=0.02). Figure 2 shows the four work groups based on not modifying work tasks or having worries (unaffected), worry about productivity (worries), modifying work tasks (tasks), and both modifying work tasks and productivity worries (both) by symptom classes. Those in the severe, GI, and mild symptom classes were more likely to have to modify their work tasks than those in the allergy group (Figure 2).

**Figure 2.**
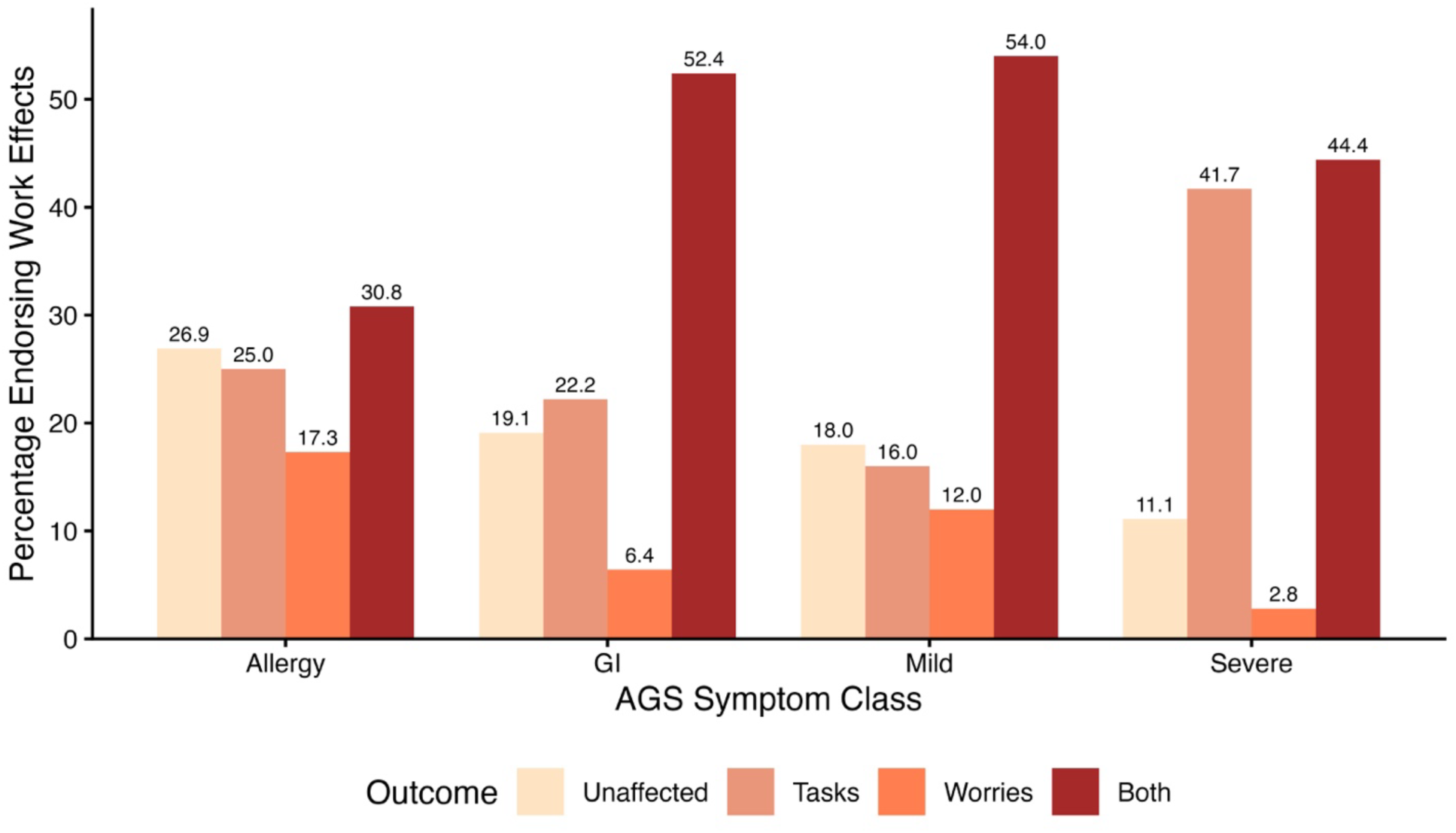
Impact of AGS on work by symptom classes in 201 agricultural operators with an AGS diagnosis, 2025. Unaffected: not modifying work tasks or having worries; Tasks: modifying work tasks; Worries: worry about productivity; Both: modifying work tasks and productivity worries

Class was associated with feeling hopeless (p=0.001) and anxiety (p=0.045), but not with being distressed (p=0.67) or depressed (p=0.06). Figure 3 shows the mental wellbeing factors by AGS symptom group. The severe AGS group shows the highest impact on all mental wellbeing indicators. Hopelessness was lowest among all the indicators and anxiety and distress were the highest across all classes (Figure 3).

**Figure 3.**
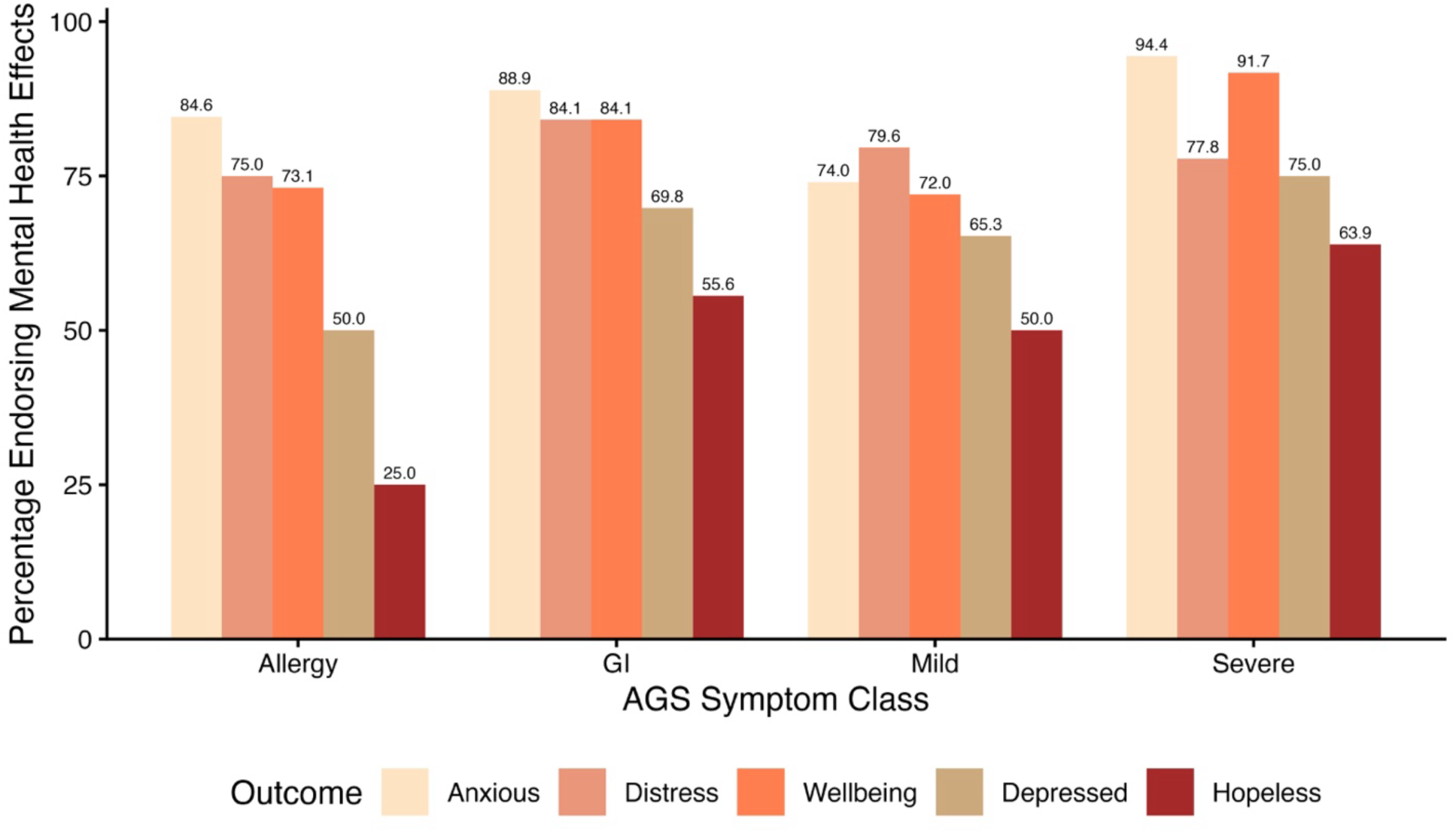
Mental wellbeing factors by AGS symptom classes in 201 agricultural operators with an AGS diagnosis, 2025.

### Qualitative Sample

The agrarian imperative can be found in the common themes and subthemes expressed by participants. The connection to the land, sharing of the heavy workload by all family members, being self-sufficient by growing the family’s food, managing the workload required by agricultural production, and the farm as a family legacy are aspects of the agrarian imperative and connects farmers to the land and animals they value. The following are major themes identified in this cohort.

#### Theme 1: Participants reported that they knew they had high exposure to ticks but it was not going to stop them from being out on their land and doing the work

Farmers and ranchers expressed their concern about tick exposures while working on their land. Eleven of the fourteen participants described finding ticks on themselves frequently when outdoors. Work tasks frequently mentioned as an exposure risk to ticks and tick bites included fencing and walking through the brush. Most participants have continued with their outdoor work responsibilities, although encounters could lead to additional symptoms.

> *“… I’ve been bit by probably 5 or 6 ticks since the diagnosis. I swell up and get a bad sore…I’m gonna go to our farm, I’m gonna do the things. I stay out of the wooded areas. They still seem to find me, but I got bit two weeks ago. … I was down there cutting, and I ended up with a tick stuck in my leg. It swelled up, it’s incredibly frustrating, because I’m trying to get ahead of the ticks before they come out, but they just seem to be everywhere.”(Farmer and Rancher, Aged 41-45, Nebraska)*
>
> *“I know that if I get bit again by a tick …that it could make it worse. I guess that is something that makes me super anxious. I’m not gonna stop going outside.” (Famer and Rancher, Aged 26-30, Missouri)*

#### Theme 2: AGS puts additional burdens on other members of the family and work tasks

Participants were forthcoming about the impacts their AGS diagnosis had on their family and work tasks. The participants reported having to delegate their tasks to other family members to avoid exposure to mammals and many mammalian fluids.

> *“I definitely do not help out with the cows unless it’s just one of those situations where it’s a life or death situation with a farm animal. I just have to stay back and hope that somebody else is here to help with that, so I know the kids have had a lot more on their plate, just from mom not helping with chores.” (Farmer, Aged 41-45, Missouri)*

Many participants were unable to completely avoid mammalian encounters on their operation, especially in emergency situations. Small family operations may be nonfunctional without the member with an AGS diagnosis being able to work. Participants forced to work with mammals to support their families often experienced symptoms following the encounter.

> *“I don’t avoid cattle. I can’t avoid cattle. It’s me and my husband that do all the work. I do notice that I just wormed 3 bulls the other day and I got kinda itchy messing with them, but we had to vaccinate some calves. I can’t stop doing. If that makes me swell up for a while, then I’m just going to swell up for a while. As far as work goes, I hurt, like my body hurts all the time. I’m not doing my job like I used to do it. Brain fog, brain fog is huge. That’s a huge problem. … I just can’t get my words out, it doesn’t just affect my body as far as physically, it affects me mentally too. I get up every day and I go to my job as best I can. There’s days I just absolutely feel like crud and I just drag and I’m sure my employees are like what is up with you. It puts a toll on your body.“ (Rancher, Aged 46-50, Missouri)*

Interestingly, some participants who avoided their mammals completely, reflected on changes to their operation including animal behavior which may have economic implications.

> *“Can’t be around the animals when they’re born to get them used to people. I guess that bothers me because they’ve gotta be nice to people if you’re gonna sell them or use them for carting. I can’t sit out there with them and be around them anymore, so their use is not the same as it used to be. Their value is no longer the same. It’s a total different adjustment.” (Farmer and Rancher, Aged 71-75, Tennessee)*

#### Theme 3: AGS means that the meat grown on the operation can no longer be eaten by one or more family members, or sometimes even be present in the household

Participants, specifically those in ranching, lamented about the drastic change to their diet. They rely on their occupation as not only a livelihood, but also as a predominant food source. Many wondered what the future of their operation will look like if they cannot eat mammalian meat.

One participant even expressed frustration over the inability to support the cattle and beef industry as they once did.

> *“We’ve changed the major proteins that we eat, which is really unfortunate because as beef farmers, we want people to eat beef, and then we’re not even supporting it now. My entire family, in order to accommodate me and my nephew, we don’t eat like near the amount of steak and burgers that we used to. …With cattle, it’s really unfortunate because we’ve raised cattle our entire life. We raise sheep now, and these are all things I can’t eat. …That was one of the big things that we raised it so that we could have food security and make sure we had what we needed. We’re still going to do cows because we all love cows, but we’re looking into potentially getting into the duck business for a little bit to, to have some alternative meats... I’m trying to potentially also get a hold of some of those alpha gal-free pigs and see if I can raise them. Not only myself, but to help raise these pigs for other alpha gal people because we are farmers. I want to put my knowledge to use.” (Rancher, Aged 31-35, Tennessee)*
>
> *“And the idea of investing money to put pens in for cattle, it’s gonna break my husband apart when I tell him ain’t gonna happen then. Why should I stay on cattle when I can’t eat it? I don’t even know how allergic I’ll be if I’ll be fume reactive to the smell of the cow. I don’t know how I’m going to have that conversation with him. You know, the sheep, all of it, I’m allergic to all of it. I thought it was just eating it, but getting the fume reaction to gelatins. What else am I gonna react to? I live out in the country, this is what I moved here for.” (Farmer and Rancher, Aged 66-70, Missouri)*

#### Theme 4: Additional financial costs of AGS when economics are already challenging

Only one participant reported hiring outside help because of their AGS diagnosis. Many other operators relied on other family members, as seen in Theme 2, or continued working through their symptoms to avoid the added expense of hired labor. Costs of operating farms and ranches have increased drastically in recent years, but AGS introduces unique financial challenges.

> *“We had to hire a farm help because to clean stalls. I have to wear gloves if I go out to help worm. The birthing is a problem because of all the fluids. I don’t clean out stalls anymore. We hired to help because I just couldn’t do it anymore*. … *Having to hire farm help to keep our farm going, and take care of the animals, that cost went up. It was not very profitable to raise and sell animals anyways. I won’t say doubled but at least added. … The farm cost has gone up about 75%, not quite 100.” (Farmer and Rancher, Aged 71-75, Tennessee)*

Outside of the operation, the cost of shopping for food, medications, and personal care products increases. People with AGS must carefully read all labels, and after their diagnosis often replace personal care and other household products. As outlined in Theme 3, these participants were used to eating meat produced on their own land. Now, many have had to find alternative sources of protein for their daily diets, which is financially burdensome.

> *“I have to stay in the kitchen a lot more cooking because I can’t eat normal stuff and so our grocery bill’s gone up for sure. I do eat ostrich and emu now when I’m craving a hamburger. … it’s cost us a lot more money. Not only food, because I have to eat something like ostrich, which is $19 a pound. I have to eat a lot of fish, which is $15 a pound. The food cost has gone way up. My food cost is more than doubled.” (Farmer and Rancher, Aged 71-75, Tennessee)*

AGS is a drastic lifestyle change for farmers and ranchers, especially given that their occupations place them primarily outdoors and around livestock. Some producers cannot continue working due to the severity of symptoms, which can be devastating for a small farming operation.

> *“I don’t think I’ve gotten a paycheck in a year and a half. When you talk about financial impact, not having an income has been the biggest thing.” (Farmer and Rancher, Aged 56-60, Oklahoma)*

#### Theme 5: Mental health impacts

The most common theme regarding the mental health impacts of AGS is anxiety around eating. The participants reported being stressed by not knowing what was safe for them to eat, especially when eating away from home.

> *“And now I’m terrified that anything else I cook is mammal-based it’s gonna make me sick. It’s bad!… Oh, I’m depressed. I don’t even feel like getting out of bed. But I have to just realize that I can eat chicken! … Should the signs of depression be addressed during the treatment of Alpha-gal? You betcha” (Farmer and Rancher, Aged 66-70, Missouri)*
>
> *“About once a week. It’s more of a breakdown of not being able to eat anything. Food has become an aversion. My daughter and I talk about this all the time. You think about all the things you’d like to eat or drink like a milkshake or regular a donut or just regular bread. It’s like once a week, it’s especially if I haven’t been grocery shopping and I’m getting low on the things I can eat. Our refrigerator is pretty bare. That’s the most stressful part.” (Rancher, Aged 51-55, Oklahoma)*
>
> *“I gotta keep control of my mouth if somebody brings something in or just makes a mistake and I end up getting sick. You gotta keep from lashing out. There’s a constant level of stress with it. Just having to be mindful all the time of what you’re exposed to.” (Farmer and Rancher, Aged 56-60, Oklahoma)*

Family support was important to participants as they navigated the dietary changes. There was a range of severity with the symptoms. Dietary adjustment was generally perceived as easier if the individual was not fume reactive and could cook for the family; even if having to make a separate meal for themselves.

> *It’s just my husband and I. Our kids are grown and out of the house right now. He’s very compassionate and understanding about the situation and he’s totally with us. Our food choices at home, and when he eats out, I encourage him to eat beef or pork or whatever he wants, but at home we’re strictly on the alpha gal diet. (Rancher, Aged 66-70, Kansas)*
>
> *“My family’s carnivores. My dad was in the meat business. I grew up east steak and roast and beef and pork. You betcha, and I can eat none, because it makes me real sick. They don’t understand. They’re trying, my husband’s sweetheart, but I still have to make him bacon for breakfast once in a while. You know, I put a pork chop in the oven and wear a bandana and wear a mask when I cook.” (Farmer and Rancher, Aged 66-70, Missouri)*
>
> *“It’s been pretty inconvenient. I guess we could say with cooking. Typically, I’m cooking two meals a night. All the time. We live on a farm, so we butchered two hogs, and they’re eating pork, bacon, sausage, anything you name, and I, of course, can’t have it, so it’s created a little bit of a stink there.” (Farmer, Aged 31-35, Missouri)*

The mental health burdens were not only influenced by food. There is worry about having a health crisis away from home and not always knowing where to receive medical care if they start having allergic reactions. Additional concerns about what allergens they may encounter when receiving care from providers (e.g. doctors and dentists) were present.

> *“You didn’t know why you’re breaking out, and we go to Oklahoma quite a bit. Where’s the nearest hospital where we’re staying? Where’s the nearest ER? So if in the middle of the night we gotta get up and go, I want to know where to go, and traveling to Kansas City to see my mom what if I break out while I’m driving in the middle to Kansas City, what do you do”? (Rancher, Aged 66-70, Kansas)*
>
> *“It was just a huge learning curve, but I didn’t want to go to doctors and stuff, or like get hurt or go to the dentist and do anything other than just like basic cleanings because I didn’t wanna have to fight all that. I didn’t wanna have to deal with it. Honestly, it was so much stress.” (Rancher, Aged 31-35, Tennessee)*

Possibly, AGS can initiate depression in those otherwise not depressed. Whether this is related to symptoms of intense fatigue or brain fog, which may mimic depression-associated symptoms, or if AGS causes neurological effects that result in depression is unknown.

> *“My wife thought I was depressed. I’m sure symptoms are different for everyone, but when I didn’t know what it was… I’m a go-getter and working and doing lots of… I’m a very hard worker. It just wiped me out. I was just tired, I had no energy. I remember on a Sunday, I just was laying around, and she was like, come on, get up, you need to get up and move, you can’t just lay around all weekend. I’m like, I’m tired. I wasn’t sick, I’m just tired. She kind of gave me a hard time about, I mean, she didn’t say the word, but I was like, why are you so lazy all of a sudden? I’m like, gosh, why am I so lazy all of a sudden? She told me later that she just thought I was depressed, because I wasn’t myself. When I finally got the diagnosis, I was actually relieved …* “ *(Farmer and Rancher, Aged 41-45, Nebraska)*
>
> *“I am stressed out all the time. I don’t know if it’s that or what it is, but yeah, constantly, and constantly I am bitchy. I get moody, and I mean, I didn’t used to be quite like that, you know, that bad. Emotional, I’m super emotional now. …Is that just my age? Is that hormones? I don’t know. My husband can probably be the better answer of that one, but my mood has changed. I have a different outlook on life. I think I’ve made myself a little bit tougher, my empathy level has kind of left a little bit.” (Rancher, Aged 46-50, Missouri)*

### Mixed Methods

The qualitative interview data reflected much of what was asked in the quantitative survey by design. However, the interview questions were general in nature to allow participants to describe their experiences with their diagnosis and to express what the experience meant to their lives. Table 3 represents an alignment between these two methodologies to demonstrate how the personal experiences inform the quantitative findings.

**Table 3.**
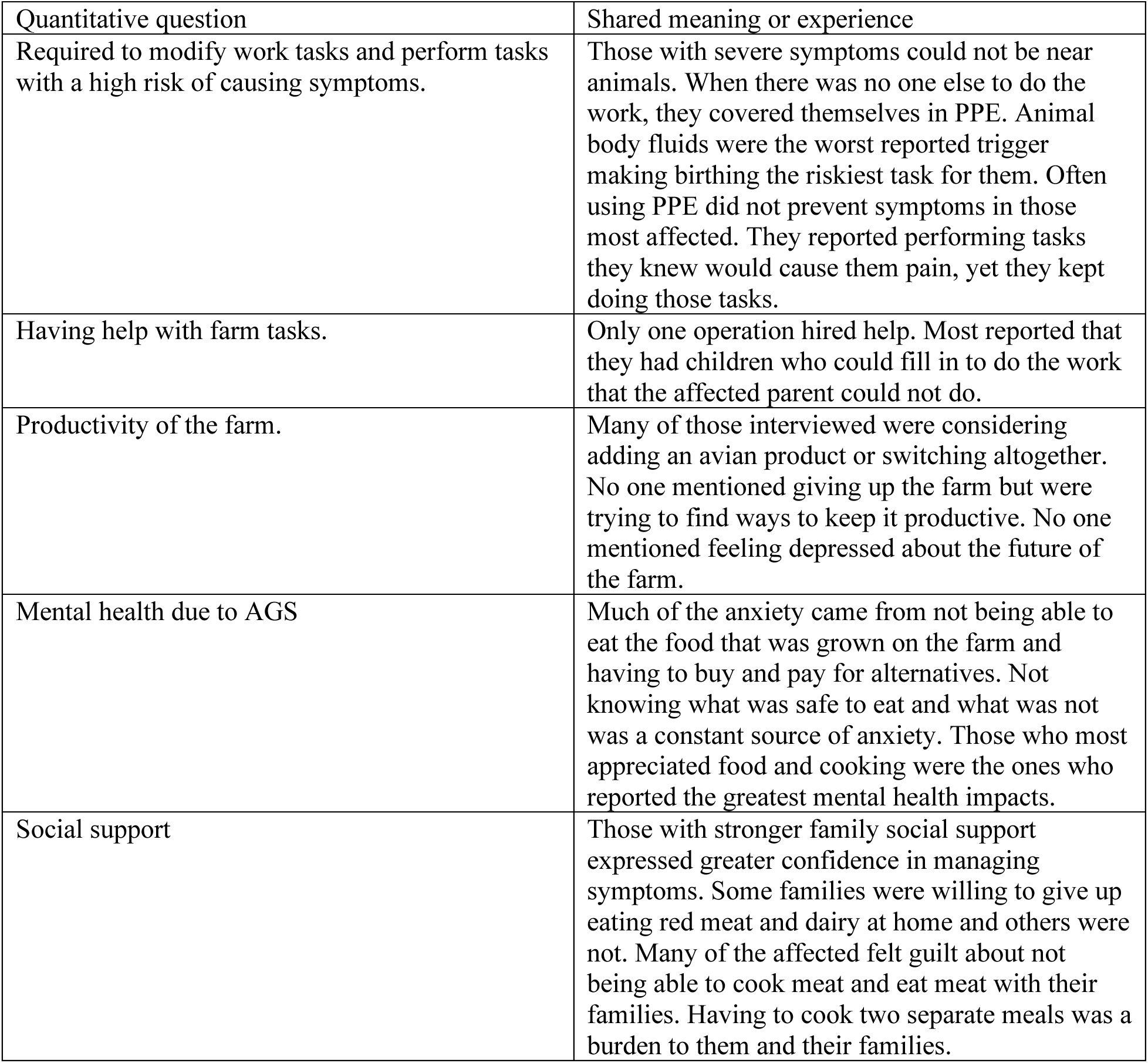
Mixed methods: Summary of qualitative analysis related to quantitative analysis.

| Quantitative question | Shared meaning or experience |
| --- | --- |
| Required to modify work tasks and perform tasks with a high risk of causing symptoms. | Those with severe symptoms could not be near animals. When there was no one else to do the work, they covered themselves in PPE. Animal body fluids were the worst reported trigger making birthing the riskiest task for them. Often using PPE did not prevent symptoms in those most affected. They reported performing tasks they knew would cause them pain, yet they kept doing those tasks. |
| Having help with farm tasks. | Only one operation hired help. Most reported that they had children who could fill in to do the work that the affected parent could not do. |
| Productivity of the farm. | Many of those interviewed were considering adding an avian product or switching altogether. No one mentioned giving up the farm but were trying to find ways to keep it productive. No one mentioned feeling depressed about the future of the farm. |
| Mental health due to AGS | Much of the anxiety came from not being able to eat the food that was grown on the farm and having to buy and pay for alternatives. Not knowing what was safe to eat and what was not was a constant source of anxiety. Those who most appreciated food and cooking were the ones who reported the greatest mental health impacts. |
| Social support | Those with stronger family social support expressed greater confidence in managing symptoms. Some families were willing to give up eating red meat and dairy at home and others were not. Many of the affected felt guilt about not being able to cook meat and eat meat with their families. Having to cook two separate meals was a burden to them and their families. |

## Discussion

### The Agrarian Imperative and Alpha-gal Syndrome

The agrarian imperative posits that people have an innate compulsion to connect to the land and will sacrifice to maintain that connection [8]. In this study, participants reported even though AGS increased risk in work tasks, this was not going to stop them from being out on their land and completing their work. Not a single participant mentioned selling their operation.

Participants had a variety of perceptions of the risk of future tick bites (Theme 1). Some chose to take extreme protective measures while others felt comfortable just checking for ticks. They stated they would not let a tick bite keep them from doing the work needed to be done or from being outdoors on their land. They modified their work to avoid ticks when possible but most accepted it as part of farm life. The worry that comes with additional tick bites likely caused a degree of stress, which they learned to manage.

Family participation in the work of the agricultural operation is inherent in farming culture. It is expected that everyone will do age-appropriate tasks. AGS puts additional burdens on unaffected family members (Theme 2). Participants acknowledged the burden that they put on family members when their symptoms were of a magnitude that they could not work on the operation. Most of them chose to wear all the protective equipment available to them and do the best they can with preventing contact with animals when forced to perform risky tasks. They knew there were times when they had to risk becoming ill because there was no one else to complete the work.

A driving force for the agrarian imperative is self-sufficiency. Furthermore, maintaining a productive farm was the most important objective among participants in our interviews. Yet, AGS causes stressors on farmers and ranchers that challenge this self-sufficiency (Table 3; Themes 3 and 4). AGS means the meat grown on the operation can no longer be eaten by one or more family members, or sometimes even be present in the household (Theme 3). Nearly every participant in the interviews mentioned not being able to eat what was grown on their operation and the added expense of having to buy food they historically did not have to purchase. This food can cause financial burdens as it often is more expensive because it is an exotic alternative or genetically modified to be alpha-gal free (Theme 4). Other participants reported the decline in income or complete job loss, both on and off the farm, due to sensitivity of AGS triggers (Theme 4). These highlight additional burdens and costs exacted by AGS due to lifestyle changes; exacerbating pressures already present in agricultural operations.

The distress experienced by participants is likely a combination of working the land to grow their own food yet unable to reap the benefits of the work and trying to keep the operation productive with forced work modifications. Several participants were considering changing to avian products such as chickens, emus, and ostriches to grow alpha-gal safe food. The ranchers that were attached to beef cattle struggled the most with the diagnosis based on our qualitative assessments.

### Mental health impacts

Participants reported high levels of anxiety, distress, depression, and a lower sense of wellbeing (Table 1, Figure 1; Theme 5). They were less frequently hopeless. Their experiences fit the Lazarus Transactional Model of Stress and Coping [22,23], which recognizes that a persistent misalignment between perceived environmental demands and an individual’s available resources manifests in distress. In this model, a stressor is appraised for its potential threat level, and the requisite coping resources and coping responses lead to short- and long-term outcomes. An acute stressor over time becomes chronic distress.

Distress was related to having to modify work tasks, perform risky tasks, and productivity worries, which are ongoing concerns with persistent symptoms (Figure 1, Table 2). This was stark with 45.8% reporting distress, and individuals were 18 times more likely to experience distress when they modified work tasks and have worries over the operation’s productivity. Social support did not reduce distress. Anxiety, depression, social withdrawal, and reduced functioning can result from long-term distress [22,23].

Participants experienced assessing the threat of their AGS diagnosis in terms of impacts to their agricultural operation and lifestyle. Those who were interviewed did not explicitly state that AGS-related work issues caused them stress. They adapted to AGS via drastic work and dietary changes, but many reported mental health consequences after the diagnosis (Theme 5). The constant worry about being made ill by doing farmwork would be a long-term stressor leading to being in a state of distress, mitigated by coping mechanisms to manage the distress. Since many of the participants were still feeling this distress years after the diagnosis, they likely are in a chronic distress mode and using whatever coping skills they have available to them.

Food-related anxiety was experienced by 93% of the participants in the quantitative study (Table 1). In interviews, food was the factor most discussed and labeled as “stressful” and “anxiety-inducing.” Two primary challenges were observed from the qualitative data. The first was not knowing what was safe to eat. Participants reported a learning curve with understanding what food could cause symptoms and what would not. A predominant factor in food-associated stress was that AGS feels like a moving target. Sometimes what food they assumed would be a problem was not and what they thought was safe for them to eat caused symptomatic reactions. The second challenge was the fear of being away from home and worrying about where they could receive medical care if they had severe symptoms. Eating outside the home and traveling were stressful situations for participants.

In studies of mood disorders in agricultural operators, women are found to have greater anxiety and depression than men [24,25]. This is often due to women filling a variety of roles by being the primary caregiver and working both on and off the farm [26]. We observed women reporting greater anxiety than men. However, surprisingly in this study, men and women were equally likely to feel depressed and men were more likely to seek behavioral healthcare. It is also unusual to find that men sought behavioral health support more often as this is atypical compared to other studies of mental health among male agricultural workers [27–31].

The phenotype classes identified in the previous study [15] were supported by this current study. All groups reported task modifications; however task modification was more often observed in the severe, gastrointestinal (GI), and mild symptom groups. The more severe groups showed more hopelessness and anxiety compared to the less severe groups (Figure 3). This indicates severity of symptoms is likely important in managing the work tasks. Among our fourteen interviewees, seven of the participants were in the GI symptom group, three were each in the allergy and mild symptom groups, and only one person was in the severe symptom group. Of these participants, five of the seven GI group members, two of the three allergy group members, and all mild and severe group members reported work modifications for a total of eleven of fourteen reporting modifications. Given the low number of participants interviewed with severe symptoms, it is likely that our results underestimate the impacts of AGS on farmers and ranchers by phenotype classification alone.

This study has several limitations that should be considered. The respondents in both the quantitative and qualitative study were mostly female and the majority were animal producers. This may represent those who are most affected because those most affected have greater reason to respond to the survey. The sample may not fully represent male experiences and those who are primarily crop farmers but who might keep a few animals. The source of the AGS respondents is also a factor. In the U.S. there are more females using Facebook than males. However, a separate national patient-survey sample of AGS shows a comparable pattern of skewed response rates by females (81.5% female) [32]. The quantitative study should be repeated in a larger study of agricultural operators and farm workers. This study did not cover migrant, immigrant, or seasonal farmworkers who are also likely to be exposed to ticks and develop AGS.

## Conclusion

This study used a mixed method approach to understand the impact of AGS diagnosis on work practices and behavioral wellness using the framework on the agrarian imperative. We found that modifying work tasks was associated productivity worries and several adverse mental health outcomes. Anxiety was the most frequently reported mental health impact of AGS, but distress was more highly associated with work factors. Additionally, building on our previous research, we found symptom class membership was associated with having to modify work tasks and feeling hopeless and anxious Finally, we identified five unique themes from qualitative interviews. In line with the agrarian imperative, although AGS increased risk in work tasks, it was not going to stop participants from using their land and maintaining that connection.

## Data Availability

All data produced in the present study are available upon reasonable request to the authors

## Statements and Declarations

## Acknowledgements

We would like to thank Sharon Forsyth (Alpha-gal Alliance) for her assistance with access and promotion to community groups for recruitment to this study. We sincerely thank all the farmer and rancher participants who provided their invaluable responses and assisted in this work.

